# Hormone Replacement Therapy and Cognitive Trajectories in Females: A Longitudinal Study of Estrogen-Only and Estrogen-Progestin Therapies

**DOI:** 10.64898/2026.11.24.26368883

**Authors:** Hannah Leistle, Mahsa Dadar, Cassandra Morrison

**Affiliations:** Department of Psychology, Carleton University, Ottawa, ON, K1S 5B6, Canada; Department of Psychiatry, McGill University, Montreal, Quebec, H3A 1A1, Canada; Douglas Mental Health University Institute, Montreal, Quebec, H4H 1R3, Canada

**Keywords:** Alzheimer’s disease, cognition, cognitive decline, menopause, hormone-replacement therapy, females, estrogen, estrogen-progestin

## Abstract

**BACKGROUND:** Estrogen is thought to be neuroprotective; however, estrogen drops during menopause. Hormone-replacement therapy (HRT) could prevent cognitive decline in females by replacing endogenous estrogen. However, research has generated contradictory findings on whether HRT benefits cognition in females. The current study explores the influence of estrogen-only HRT and estrogen-progestin HRT on cognitive performance in females over time.

**METHODS:** Longitudinal data from 22,706 female participants aged 55+ (*n* = 1336 estrogen-only HRT, *n* = 144 estrogen-progestin HRT, *n* = 21,226 no HRT) was included from the National Alzheimer’s Coordinating Center. Linear mixed-effect models examined the relationships between HRT conditions and cognitive functioning over time.

**RESULTS:** Both types of HRT were associated with slower decline in cognition, with combination HRT predicting better cognitive outcomes than estrogen-only HRT.

**DISCUSSION:** The findings suggest that HRT may offer protective effects for cognition over time, especially estrogen-progestin HRT, contrary to some previous research; however, more research is needed.

## 1. Background

As the proportion of older adults increases, so does the number of people affected by dementia, straining families and the health system [1]. Dementia rates differ by sex: females are 50% more likely to develop dementia than males, accounting for 66% of Alzheimer’s disease (AD) cases [2]. These sex differences highlight the need for research into strategies to prevent female cognitive decline.

Initial research on sex disparities in dementia rates suggested that the difference was associated with females living longer than males, age being one of the strongest risk factors for dementia [3]. However, even when controlling for age, females still exhibit an increased risk of AD [4]. Another hypothesis suggests that decreasing estrogen levels in females during menopause may heighten risk. Endogenous estrogen has neuroprotective effects, is associated with neuroplasticity, and may inhibit the accumulation of beta-amyloids [5]. Thus, the decrease in estrogen during menopause could influence cognitive decline and vulnerability to AD [6].

Menopause timing also typically overlaps with the prodromal phase of AD [7]. One route to mitigate cognitive decline may be to reduce estrogen loss. As such, hormone-replacement therapy (HRT) could help prevent decline and reduce AD risk in females.

However, research on HRT and cognition in females has yielded widely varying results. The Women’s Health Initiative Memory Study (WHIMS), a randomized controlled trial (RCT) from 2003, found that females who received combined estrogen-progestin HRT were twice as likely to develop dementia compared to those who did not take HRT [8]. These results contradicted the prevailing scientific opinion of HRT being neuroprotective [7,9]. Since 2014, research on this topic has been increasing [9] but systematic reviews and meta-analysis continue to highlight conflicting results: some conclude that HRT prevents cognitive decline [10,11], others that it worsens decline [12,13], and some that the evidence is inconclusive [14,15]. This divergence of opinion applies both when the participants are healthy female adults [11–13,15] and when they are diagnosed with conditions such as AD [10,14,15].

There are several possible reasons for these conflicting findings. As many authors note [10,14,15], there is methodological heterogeneity across HRT studies. Experiments are more likely to find that HRT has a neutral or negative effect, and correlational studies more often find the opposite [7]. While experiments establish causation, they tend to be shorter in duration, with a median of only about six months [13], and HRT interventions of less than one year cannot establish increased risk [16]. Older RCTs have also had smaller sample sizes [17–19]. Longitudinal correlational research, such as the present study, takes into account the development of cognitive decline, capturing progression from healthy to pathological aging.

The other purpose of this paper is to differentiate between estrogen-only HRT and combination estrogen-progestin HRT, which includes both estrogen and progestin (synthetic progesterone). Longitudinal research, especially older studies, do not specify what kind of HRT participants received [20–23]. Recent studies also include HRT as a binary variable: one longitudinal study did not distinguish between HRT types [24], and another study excluded combination HRT users [25]. HRT formulation may be an important variable, as estrogen-only therapy appears more likely to be protective than combination therapy [9,12,15,26,27]. However, only a few longitudinal studies have examined both types at once [27–30]. These range in sample size from 493 to 2304; more and larger-scale research is still needed. Furthermore, these studies, which range in follow-up time from two to seven years, found conflicting results, such that estrogen-only HRT may be beneficial while combination HRT was detrimental [27], both types may be benign [28,30], or both detrimental [29].

The current study helps clarify the relationships between the two types of HRT and cognition. Using secondary data from the NACC, a longitudinal analysis with a large sample size to examine the differential impacts of estrogen-only and combination therapy (estrogen-progestin) on cognitive and functional outcomes was conducted.

## 2. Methods

### 2.1 National Alzheimer’s Coordinating Center participants

The data used were from the Uniform Data Set collected and provided by the National Alzheimer’s Coordinating Center (NACC). The data were collected between 2005 and 2024. Only female participants over the age of 55 were included. Variables used for the analyses are described below; the full definitions are available in the NACC Data Dictionary (NACC, https://naccdata.org/)

Cognitive status of each participant was calculated combining information from three NACC variables: NACCTMCI, indicating cognitive impairment status and type; NACCETPR, the participant’s primary, official diagnosis; and CDRSUM, the sum of the scores for the eight domains of the Clinical Dementia Rating – Sum of Boxes (CDR-SB) scale used to global cognitive status. First participants were excluded if they did not have either an official AD or Mild Cognitive Impairment (MCI) diagnosis or normal cognition (NC). The criteria for our diagnostic variable were defined as follows: participants with a CDRSUM score greater than 4.0 were categorized as having AD, those with a score between 0.5 and 4.0 as MCI, and those with a CDRSUM score of 0 as NC. Participants with other neurological disorders (Parkinson’s disease, Lewy body dementia, frontotemporal dementia, etc.) or inconsistent CDR-SB scores and diagnostic labels (e.g., individuals with CDR-SB > 4.5 and no diagnosis identified based on NACCETPR) were excluded, based on the NACC [31,32]. This method has been used in previous publications [33].

### 2.2 Hormone replacement therapies

Two binary variables were used to assess participants’ use of hormone replacement therapy: NACCEMD and NACCEPMD. NACCEMD measures whether a participant used estrogen-only hormone therapy at a given point in time, such as estradiol, estropipate or quinestrol, excluding any topical medications. NACCEPMD assesses the use of combination therapy, that is estrogen combined with progestin, in much the same way; example medications include drospirenone-estradiol and estradiol-norethindrone. Participants were classified as estrogen-HRT when they reported use of either estrogen only at baseline, and as or combined-HRT when they reported estrogen + progestin therapy at baseline. Participants were classified as no-HRT when they did not report use of either HRT.

### 2.3 Outcomes

Multiple outcome variables were included to assess changes in cognition and daily functioning. Cognitive outcomes included CDR-SB scores as a measure of global cognitive status, Functional Activity Questionnaire (FAQ) as a measure of functional status, Trail Making Test and Digit Symbol substitution tests as measures of executive functioning, animals and vegetable fluency as a measure of language, Digit Forward and Backward as measures of short term memory, and Mini-Mental State Examination as a measure of global cognition.

### 2.4 Statistical analysis

All analyses were performed using R version 4.5.3. Demographic and clinical characteristics were analyzed using independent sample t-tests and chi-square tests.

To explore the relationship between hormone replacement therapy and cognition over time a series of linear mixed effects models, adjusting for age at baseline, education, diagnostic status, and time from baseline were completed. Participant ID was included as a categorical random effect. The interaction of interest was Hormone Replacement Therapy : Time From Baseline to explore whether HRT influenced cognitive decline. No-HRT was used as the reference group in the first analysis, with a second analysis completed with estrogen-HRT as the reference group to compare estrogen only and combined therapy. Separate analyses were completed for each cognitive outcome (CDR-SB, FAQ, Trails A & B, Digit Span forward and backward, MMSE, WAIS, and Animals and Vegetable fluency).

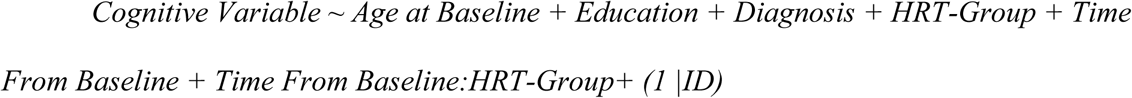

All continuous variables were z-scored prior to analyses. All p-values are reported as corrected values with significance determined by false discovery rate (FDR) controlling method to account for the multiple comparisons performed [34]. Baseline demographic differences were assessed using Levene’s tests for homogeneity of variance. When this assumption was violated, Welch’s ANOVA followed by Games–Howell post hoc tests was used.

Finally, due to the difference in age between groups, age-matched supplementary analyses were conducted.

## 3. Results

### 3.1 Demographics

After the exclusions described above, a total of 22,706 participants remained (*n* = 1336 only-estrogen HRT, *n* = 144 combination HRT, *n* = 21,226 no-HRT). Demographic and clinical statistics for all participants are presented in Table 1.

**Table 1:** Demographic and clinical characteristics for participants.

|  | **Estrogen-only HRT** | **Combination HRT** | **No HRT** |
|---|---|---|---|
| Total at baseline | 1336 | 144 | 21226 |
| Age | 69.99 ± 8.74 | 65.96 ± 7.61 | 72.50 ± 9.17 |
| Education | 15.69 ± 2.83 | 15.88 ± 2.94 | 14.73 ± 3.42 |
| Baseline diagnostic status |  |  |  |
| NC | 705 (52.77%) | 82 (56.94%) | 8234 (38.79%) |
| MCI | 484 (36.23%) | 42 (29.17%) | 8285 (39.03%) |
| AD | 147 (11.00%) | 20 (13.89%) | 4707 (22.18%) |
| Follow-up years | 3.37 ± 3.98 | 4.12 ± 4.44 | 2.83 ± 3.60 |

*Notes:* NC = Normal Controls; MCI = Mild Cognitive Impairment; AD = Alzheimer’s Disease. Combination = Estrogen-Progestin. Values are presented as mean and standard deviation or total number in sample and percentage of sample.

There were significant age differences between all three HRT groups (all *p’*s < .001). Participants who did not take HRT were the oldest (*M_age_* = 72.50), followed by estrogen-only users (*M*_age_ = 69.99), and then the combination therapy group (*M_age_* = 65.96).

Significant differences between participants’ education at baseline were also observed. Participants who did not take HRT had less education (*M_educ_* = 14.73) than the estrogen-only group (*M_educ_* =15.69; *p* < .001), and the combination therapy group (*M_educ_* = 15.88; *p* < .001). The difference between HRT groups was not significant (*p* = .74).

Baseline diagnostic status also differed (χ^2^ = 155.6, *p* < .001) between the groups. While there was no difference between the two HRT groups (*p* = .60), participants who did not take HRT differed from both HRT groups (all *p*s < .001). Compared to the HRT groups, the group who did not use any HRT had a higher proportion of AD (χ^2^ = 98.00, *p* < .001) and MCI (χ² = 9.83, *p* = .007), with a smaller proportion of cognitively normal participants (χ^2^ = 120.48, *p* < .001).

### 3.2 HRT and Cognition Outcomes

Table 2 presents the Time From Baseline by Group interaction estimates from the linear mixed effects modeling examining the influence of HRT on cognitive decline. The statistically significant results are presented graphically in Figures 1 and 2.

**Figure 1:**
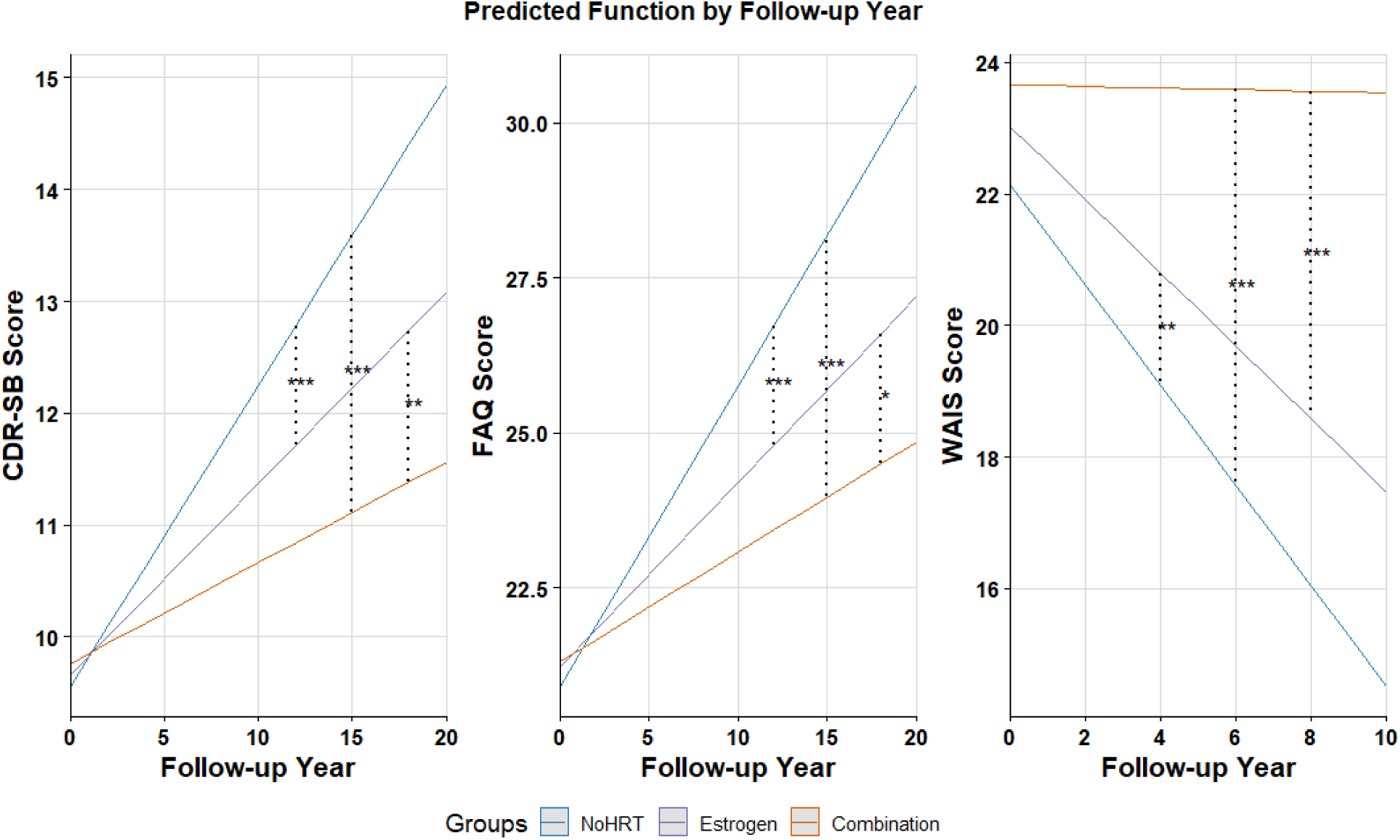
Predicted CDR, FAQ and WAIS Scores by Follow-up Year and HRT Group. *Notes:* CDR-SB = Clinical Dementia Rating - Sum of Boxes; FAQ = Functional Activities Questionnaire; WAIS = The Wechsler Adult Intelligence Scale Digit Symbol test. Asterisks represent significant pairwise comparisons. * = p ≤ .05, ** = p ≤ .01, *** = p ≤ .001.

**Figure 2:**
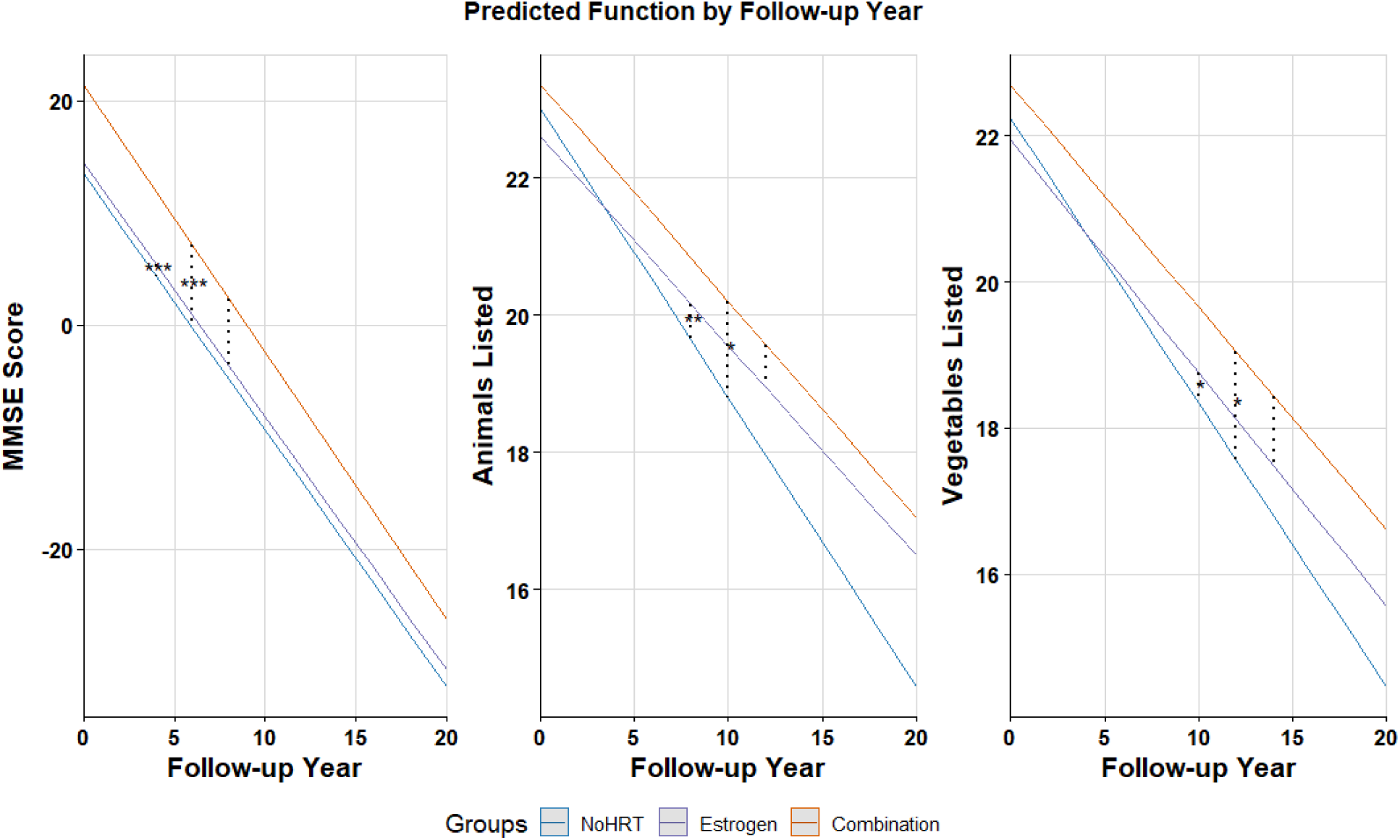
Predicted MMSE, Animal- and Vegetable-Naming Scores by Follow-up Year and HRT Group. *Notes:* MMSE = Mini Mental Status Examination. Asterisks represent significant pairwise comparisons. * = p ≤ .05, ** = p ≤ .01, *** = p ≤ .001.

**Table 2:**
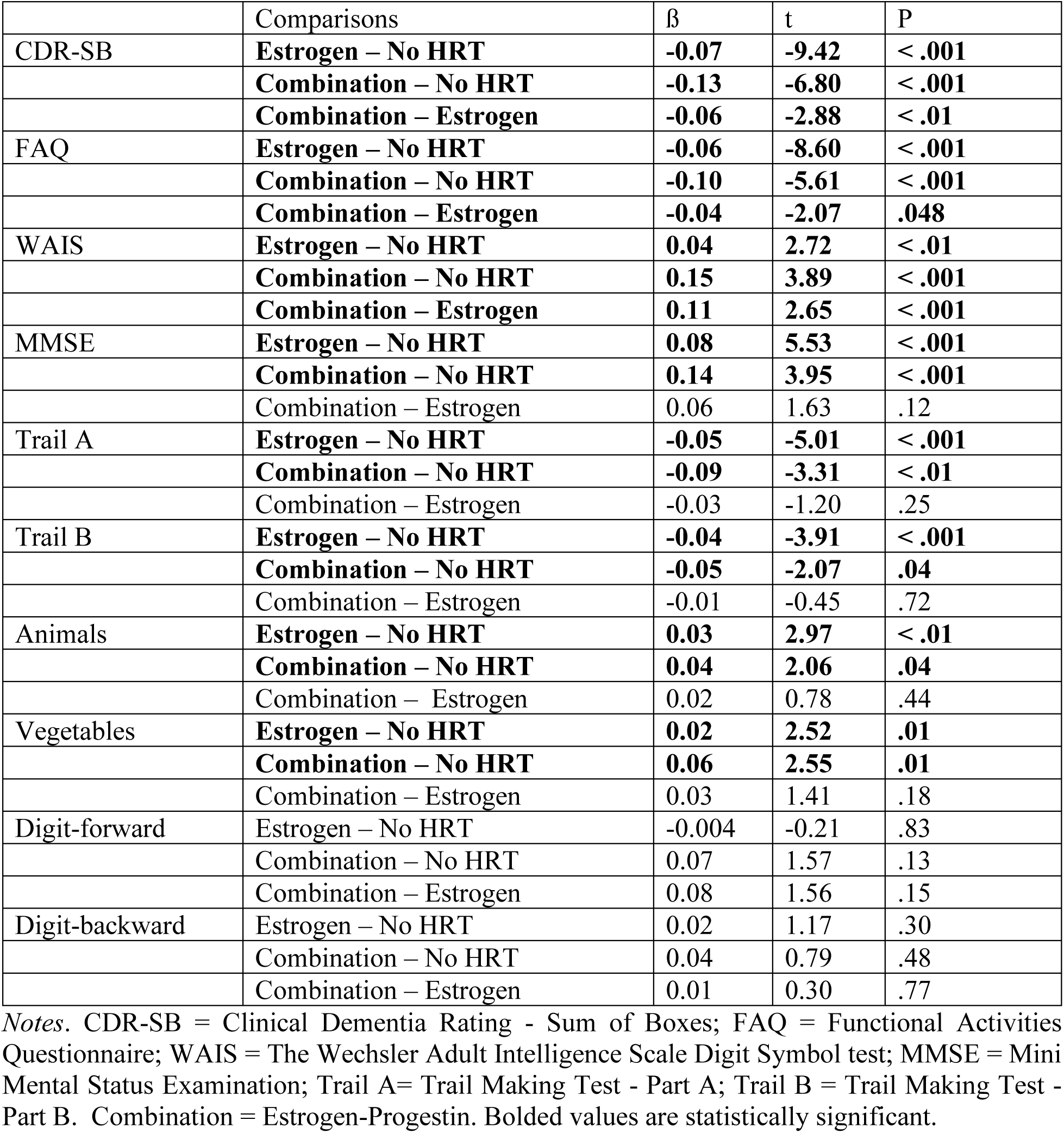
Pairwise comparisons of HRT groups and cognition outcomes by follow-up years.

Those with no hormone replacement therapy exhibited steeper cognitive decline as measured by the CDR-SB over time compared to both those who used estrogen-only HRT (*t* =9.42, *p* < .001) and those who used combination HRT (*t* = 6.80, *p* < .001). Additionally, those who used combination HRT experienced less steep declines compared to those on estrogen (*t* = 2.88, *p* < .01). Similarly, those with no hormone replacement therapy exhibited steeper functional decline as measured by the FAQ over time compared to both those who used estrogen-only HRT (*t* = 8.60, *p* < .001) and those who used combination HRT (*t* = 5.61, *p* < .001). Additionally, those who used combination HRT experienced less steep declines compared to those on estrogen (*t* = 2.07, *p* = .048). Those with no hormone replacement therapy exhibited steeper cognitive decline as measured by the MMSE over time compared to both those who used estrogen-only HRT (*t* = −5.53, *p* < .001) and those who used combination HRT (*t* = −3.95, *p* < .001). There was no significant difference between the two HRT groups (*t* = −1.63, *p* = .12).

Participants who did not take any HRT declined more in their performance on the Trail Making Test compared to both the estrogen-only and the combination HRT groups. This finding was observed for both Part A (*t* = 5.01, *p* < .001; *t* = 3.31, *p* < .01) and Part B (*t* = 3.91, *p* < .001; *t* = 2.07, *p* = .04), respectively. There was no difference in performance between the HRT groups (all *p*s > .25).

Those who did not take any HRT had lower scores in both animal (*t* = 2.97, *p* < .01) and vegetable fluency (*t* = 2.52, *p* = .01) over time compared to the estrogen-only group. Similarly, those who did not use any HRT had lower scores in both animal (*t* = 2.06, *p* = .04) and vegetable fluency (*t* = 2.55, *p* = .01) over time compared to those who used combination HRT. No differences between HRT groups were found for either fluency task (all *p*s > .18).

There were no significant differences between any of the groups over time for the digit-forward and digit-backward tests (all *t’*s < |1.58|, all *p’*s > .13). Those with no-HRT scored progressively lower on the WAIS-R Digit Symbol test over time compared to both those who used estrogen-only HRT (*t* = −2.72, *p* < .01) and those who used combination HRT (*t* = −3.89, *p* < .001). Additionally, the test scores of those who used combination HRT declined less compared to those on estrogen-only HRT (*t* = −2.65, *p* < .001).

Similar trends were observed in the results of the age-matched analyses. However, due to the small sample size, many results were no longer statistically significant. The full results of the matched analyses are provided in the supplementary materials.

## 4. Discussion

During menopause both cognition and estrogen decline [5,35,36]. Endogenous estrogen loss likely affects numerous bodily functions, including cognition [5–7]; however, results are mixed regarding the use of HRT to counter this loss and cognitive decline. This study examined whether the use of estrogen-only and combination estrogen-progestin therapy are associated with changes in cognition and daily functioning over time. The results indicate that both estrogen-only and combination HRT appear neuroprotective: participants in these groups experienced significantly less cognitive decline as measured by multiple variables, most importantly their CDR-SB and FAQ scores, compared to participants who did not use any HRT. Contrary to most of the literature, combination HRT had a more consistent beneficial association with cognition than estrogen-only HRT.

For the main outcome variables, the CDR-SB and FAQ scores, assessing global cognitive decline and daily functioning respectively, as well as the WAIS, the pattern was clear. The combination group saw the least decline, followed by the estrogen-only group, followed by the non-HRT group. All groups were significantly different from one another. Overall both types of HRT were associated with less decline, in agreement with some previous research [10,11], and combination HRT often outperformed estrogen-only HRT. These findings also agree with research suggesting that endogenous estrogen positively affects female cognition [35–41], and that a longer duration of endogenous exposure, and later onset of menopause, correlate with less cognitive decline [37].

The results for the domain-specific cognitive tests, such as the trail-making and animal-naming tests, suggest that both types of HRT may be equally beneficial for some domains. Past research confirms that HRT can have differential impacts on domains such as language functioning and short-term memory [14]. Although it would be valuable to determine if differences in cognitive trajectories are differentially associated with AD progression, there were relatively few conversions from NC to AD in the current sample, limiting our ability to explore such associations. Future research with larger samples and high rates of progression to AD are needed to further investigate this relationship.

The literature on progressive cognitive decline after menopause and the use of HRT is divided. Because HRT cannot simultaneously increase [8,12,13], decrease [10,11,42], and fail to affect [14,15,30] cognitive progression, the divergence may be due partly to methodological differences. The present results that HRT is neuroprotective align with other longitudinal studies more so than experimental designs, though it is important to note that at least one longitudinal experiment has found support for the neuroprotective effects of HRT [42]. The authors found that combination HRT helped prevent disease progression and slowed the decline of global cognitive functioning in female participants with MCI [42]. However, unlike the present study, they did not compare the two types of HRT, did not include participants who were cognitively normal at baseline, and had a sample size of 21 [42].

There are a few mechanisms through which HRT may mitigate cognitive decline. Estrogen is unique because it is produced by astrocytes and neurons in addition to the endocrine system [38], and the locations of estrogen receptors in the brain overlap with areas involved in memory and cognition [5]. Estrogen helps maintain hippocampal neuroplasticity, and may inhibit beta-amyloid formation [5,43,44]. There may also be indirect mechanisms. Both inflammation and vascular disease are linked to cognitive decline and dementia [45,46]. Estrogen reduces inflammation [47], and HRT has been used to treat patients with inflammatory conditions [48]. Similarly, estrogen helps prevent cardiovascular disease, and it protects and helps repair the heart from infarctions [49–51]. HRT can prevent age-related decline in vascular function [51]. Therefore, by preventing inflammation and vascular disease, HRT may also reduce cognitive decline. Future research should explore these pathways.

In the current study, the most surprising finding was that combination HRT had a greater number of significant associations with cognitive decline than estrogen-only HRT. While the combination group was the smallest, these participants had the longest average follow-up time, providing additional support for the observed findings. Progestin is added to an HRT formulation to reduce the risk of uterine cancer in individuals with a uterus, effectively “opposing” the negative effects of estrogen [52]. Multiple studies have found that combination HRT has a negative [29,53,54] or neutral [55] impact on female cognition. One study comparing the two found beneficial effects for estrogen-only and negative effects for combination HRT [56]. Other research studies and reviews have found that combination HRT has beneficial effects [42], but not to the same extent as estrogen-only HRT [7,57]. Only one previous study found that combination was superior to estrogen-only HRT [58]. Notably, analyses using the matched sample yielded comparable favourable cognitive outcomes between estrogen-only and estrogen-progestin HRT, providing further evidence in support of progestin.

The interaction between estrogen and progestin is complex. Progestin is added to combination HRT in order to antagonize estrogen and therefore also antagonizes some of estrogen’s neuroprotective effects [59,60]. However, there is related research that supports the protective effect of progesterone. For example, in mice, progestin prevents the beneficial effect of estrogen on beta-amyloid accumulation but not on behaviour, and prevents neurofibrillary tangles [61]. Several narrative reviews conclude that endogenous progesterone has neuroprotective effects, separately from estrogen, in mice as well as human females [59,62]. An experiment that administered estrogen and progestin separately to menopausal participants found that they activated different areas of the brain to produce different cognitive benefits [63]. The two hormones appear to have different, and at times complementary, effects on cognition and dementia that may have been overlooked in the literature.

The type of progestin may be crucial. For example, there is evidence that Medroxyprogesterone acetate, the most common kind of progestin added to combination HRT, may be harmful, whereas other kinds are neuroprotective [62]. This difference would explain some of the diverging results in the literature. Unfortunately, the NACC dataset does not specify HRT formulas, but it could be that the participants included in the present study used combination HRTs formulated to preserve the neuroprotective effects of both hormones. Future research should explore the different types of combination HRT.

The present study has a few limitations. While it assesses longitudinal change in cognition as predicted by HRT use at baseline, it remains correlational and does not establish causality. In addition, the participants who used combination therapy were fewer and younger compared to the other groups. After age matching, however, effect sizes remained largely similar; therefore the loss of significance for some comparisons may be explained by the smaller sample size.

Another limitation is that the study could not directly address the critical window hypothesis. This hypothesis posits that HRT may be beneficial for cognition when started close to menopause, but detrimental if started too late, over the age of 65 [64]. Many reviews find support for the critical window hypothesis [65–67], and the advanced age of the participants may explain the results of the WHIMS study that temporarily halted HRT research [7,14]. However, some studies with participants aged around 75 have still found protective or neutral effects for HRT [58,68,69]. The NACC dataset does not include information on when a participant started HRT. The average overall sample age was around 70, and the average person who uses HRT begins use in their early 50s, and continues use for about seven years [70]. Given that the present study found that HRT may be neuroprotective, the fact that the sample age was older would refute the critical window hypothesis, but if one assumes the participants started HRT in their 50s on average, the hypothesis holds. Without duration data, one can only speculate.

Finally, while the present study differentiated between the two broad types of HRT, it could not account for variations in formulation, dosage, and administrative routes, beyond the fact that topical methods were not included. The potential differences in impact depending on the type of progestin have already been discussed, and research has found that different types of estrogen also have different effects on female cognition, such that 17β-Estradiol may be superior to conjugated equine estrogen [71,72].

Despite these limitations, the present study is a strong addition to the literature and underscores the importance of differentiating between HRT types. Contrary to some previous research, it found that HRT, especially combination HRT, is associated with less cognitive decline compared to not using HRT. It is important that the current research momentum is maintained after the WHIMS RCT stalled its progress, given the potential benefits of HRT. Indeed, an additional strength of the present study is its naturalistic approach: participants used HRT for routine clinical care, likely symptom management, rather than through an RCT. In RCTs, participants are carefully selected, with standardized protocols. However, in the current study the females using HRT are doing so in a naturalistic real-world context, providing valuable insight into the potential positive cognitive-HRT associations that occur in everyday clinical practice. Together, these findings suggest that HRT may play an important role in mitigating cognitive decline in older adult females.

## Data Availability

All data produced are available online at https://naccdata.org/

## Declarations

## Consent Statement

Written informed consent was obtained from participants or their study partner at the institutions that collected the data.

## Consent for Publication

Not applicable

## Availability of data and materials

Data used in preparation of this article were also obtained from the National Alzheimer’s Coordinating Center (NACC; https://naccdata.org/) database,” specifically the NACC Uniform Data Set (UDS)

## Conflict of Interest

The authors declare no competing interests.

## Funding

The present study is supported by research funds from the Canadian Institutes of Health Research (CIHR). Dr. Dadar also reports receiving research funding from the Fonds de Recherche du Québec - Santé (FRQS, https://doi.org/10.69777/330750), Natural Sciences and Engineering Research Council of Canada (NSERC), and Brain Canada. Dr. Morrison reports receiving research funding from CIHR and NSERC.

## Authors’ Contributions

H.L., M.D., & C.M., were involved with study conceptualization, data analysis, data cleaning, and interpretation. H.L. & C.M. were involved with the original manuscript draft, and all authors were involved with editing and approving the final draft.

## Acknowledgments

The National Alzheimer’s Coordinating Center (NACC) database is funded by the National Institute on Aging (NIA)/National Institutes ofHealth (NIH) Grant U24 AG072122. NACC data are contributed bythe NIA-funded Alzheimer’s Disease Research Centers (ADRCs): P30AG062429 (PI James Brewer, MD, PhD), P30 AG066468 (PI OscarLopez, MD), P30 AG062421 (PI Bradley Hyman, MD, PhD), P30AG066509 (PI Thomas Grabowski, MD), P30 AG066514 (PI MarySano, PhD), P30 AG066530 (PI Helena Chui, MD), P30 AG066507 (PIMarilyn Albert, PhD), P30 AG066444 (PI David Holtzman, MD), P30AG066518 (PI Lisa Silbert, MD, MCR), P30 AG066512 (PI Thomas Wis-niewski, MD), P30 AG066462 (PI Scott Small, MD), P30 AG072979(PI David Wolk, MD), P30 AG072972 (PI Charles DeCarli, MD),P30 AG072976 (PI Andrew Saykin, PsyD), P30 AG072975 (PI JulieA. Schneider, MD, MS), P30 AG072978 (PI Ann McKee, MD), P30AG072977 (PI Robert Vassar, PhD), P30 AG066519 (PI Frank LaFerla,PhD), P30 AG062677 (PI Ronald Petersen, MD, PhD), P30 AG079280(PI Jessica Langbaum, PhD), P30 AG062422 (PI Gil Rabinovici, MD),P30 AG066511 (PI Allan Levey, MD, PhD), P30 AG072946 (PI LindaVan Eldik, PhD), P30 AG062715 (PI Sanjay Asthana, MD, FRCP), P30KAMAL ET AL . 13 of 15AG072973 (PI Russell Swerdlow, MD), P30 AG066506 (PI GlennSmith, PhD, ABPP), P30 AG066508 (PI Stephen Strittmatter, MD, PhD),P30 AG066515 (PI Victor Henderson, MD, MS), P30 AG072947 (PISuzanne Craft, PhD), P30 AG072931 (PI Henry Paulson, MD, PhD), P30AG066546 (PI Sudha Seshadri, MD), P30 AG086401 (PI Erik Roberson,MD, PhD), P30 AG086404 (PI Gary Rosenberg, MD), P20 AG068082(PI Angela Jefferson, PhD), P30 AG072958 (PI Heather Whitson, MD),and P30 AG072959 (PI James Leverenz, MD).

## Notes

### Author Declarations

Data used in preparation of this article were also obtained from the National Alzheimer's Coordinating Center (NACC; https://naccdata.org/) database, specifically the NACC Uniform Data Set (UDS)

